# Quantitative Epithermal Neutron Activation Analysis of Seven Micro Elements in Breast Milk of Lactating Mothers from the Central region of Ghana

**DOI:** 10.1101/2020.06.30.20143446

**Authors:** Irene Korkoi Aboh, Michael K. Vowotor, Andrews Adjei Druye

## Abstract

**Background:** Epithermal instrumental neutron activation analysis (EINAA) technique is used for the determination and estimation of the concentration levels of micronutrients such as Sodium (Na), Magnesium (Mg), Potassium (K), Calcium (Ca), Manganese (Mn), Copper (Cu) and Iodine (I) in breastmilk.

**Aim:** To examine the concentration of seven micro elements in the breastmilk of lactating mother who were exclusively breastfeeding.

**Methods:** This study employed quantitative experimental research where 27 lactating mothers voluntarily participated in the study from two health facilities in the Cape Coast Metropolitan area. Data was collected over a period of four weeks. A three millimetre (3mm) thick of flexible boron was used to cut off thermal neutrons in order to assess epithermal neutrons. This was done to create an activation energy which examines the amount of the 7 micro nutrients in the breastmilk. The standard reference materials used were the International Atomic Energy Agency (IAEA)-336; IAEA-407, IAEA-350 and National Institute of Standard and Technology (NIST) USA SRM 1577b. The Relative standardization method was used in the quantification of the elements.

**Results:** The study achieved about 94.7% accuracy. The estimated health risk calculated showed that the concentrations of chlorine (Cl) and iodine (I) were high in the order I > Cl with all very far above the maximum Upper Limit (UL) of the daily Recommended Dietary Allowance (RDA) for all life stages except for children below 8 years.

**Conclusion:** Mn is found in very high quantities in the diet consumed by the parents of babies. These children could be exposed to metabolic disorders or unexplained diseases in future without knowing their origin.

## Background

Breast milk is the natural first food for babies. It provides all the energy and nutrients that the infant needs for the first six months of life. Breast milk promotes sensory and cognitive development, and protects the infant against infectious and chronic diseases. Exclusive breastfeeding reduces infant mortality due to common childhood illnesses such as diarrhoea or pneumonia, and promotes a quicker recovery from illnesses (Department of Health & Human Services, (2016). Babies go through a rapid period of growth after birth. In fact, they usually double their length and triple in weight within the first twelve months of life. Even when solids foods are introduced breast milk remains an important source of nutrition for proper growth and development (Bubbahood, 2019). The Academy of Nutrition and Dietetics recommends that infants consume breast milk alone for the first six months of life, and breast milk with complementary foods from 6 to 12 months of age. Mothers who choose not to, or are unable to breastfeed can offer their baby infant formula in place of breast milk. The decision to breastfeed or formula feed a baby could be personal or influenced by health issues such as HIV/AIDS infection. Weighing the pros and cons of breastfeeding or formula feeding method can help make a good decision for the baby (KidsHealth, 2017). Infants from 0 to 6 months should take breast milk or infant formula on demand, to help meet nutritional requirements (Coleman, 2018). For instance, **carbohydrates** are the main source of energy used by the body; **Protein** contributes to a small amount of the energy used by the body and is essential for the building the tissues of the growing body. **Fat** also contributes to the energy requirements of the body, and also helps in the absorption of fat-soluble vitamins (Vitamin A, D, E and K). **Vitamins & Minerals** – are groups of nutrients which assist in protection and functioning of the body (Bubbahood, 2019).

Iodine in a trace plays an important role in human physiological actions of thyroid hormones production from the thyroid gland (Endocrineweb, 2017). Thyroid cells combine with Iodine and the amino acid tyrosine to make T3 and T4 which are released into the blood stream and are transported throughout the body where they control metabolism. The leading preventable causes of brain damage, that is Iodine deficiency can be significantly lower the intelligence Quotients (IQ) of a whole population. It has been estimated that 50 countries could prevent the loss of intellectual capacity by 10 to 15 percent points if young children, new born and pregnant mothers received enough iodine (Caulfield, Richard, Rivera, Musgrove & Black, 2006). There is the concern that the continuous occurrence of Iodine Deficiency Disorders (IDDs) among children in Ghana may hamper the objectives of the educational reform programmes and the nation’s developmental efforts. Statistics indicates that about 81,200 babies are born annually with mental impairments as a result of such deficiencies, and suffers from stunted growth and low IQs, thereby impeding their learning abilities when they grow up (Ghana News Agency, 2007). Infants up to 6 months need an iodine intake of 90 micrograms per day. Breast milk is the only best source of iodine for babies and helps in development of their brain and nervous system (Mrunal, 2018). Another important micro-mineral is magnesium (Mg) is responsible for approximately 800 enzymatic functions in the human body and one of the most important minerals that promotes the health of children and adults. Babies up to the age of 6 months may require 30 mg per day. It helps children to get better sleep to conserve energy. It also aids in insulin and blood sugar regulation, as well as DNA formation (Mahak, 2018).

Scientists have described how adequate amounts of copper are important to brain function Tiny amounts of Copper (Cu), within certain enzymes in the brain helps form key neurotransmitters that allow brain cells to communicate to one another. (US Department of Agriculture, 2007). Babies up to the age of 6 months may require 0.20 mg per day (Food and Nutrition, 2001). The normal term infant is born with a generous store of copper in the liver making copper deficiency a rare event (Widdowson, 1974). Copper deficiency might be responsible for a resistant anemia in milk-fed infants because cow’s milk is one of the few commonly used foods low in copper (Picciano, 1985). Manganese (Mn) is involved in the formation of bone (Food and Nutrition, 2001). Mn aids in the action of some enzymes involved in carbohydrate metabolism (Lenntech, 2019). Babies with either relatively high or low levels of manganese in their blood may be affect with poor brain development (Norton, 2010). Fatness, changes of hair colour, abnormal bone and cartilage function, growth retardation may result due to adverse effect of Mn deficiency (Lenntech, 2019). Babies up to the age of 6 months may require 0.003 mg of Mn per day (Food and Nutrition, 2001).

Sodium (Na) is an electrolyte/mineral that functions as a major ion of the extra cellular fluid. It also aids nerve impulse transmission. Babies to the age of 0 - 6 months may require 120.0 mg of Na per day (Health supplements nutritional guide, 2017). The document further added that the major source of sodium is from salt and before a baby is six months, baby would have had all the sodium needed from breast milk so there is no need for additional salt to baby’s food. Potassium (K) works with sodium to control the body’s water balance, which helps maintain normal blood pressure. In fact, a diet that is low in potassium and high in sodium appears to be a factor in high blood pressure. Potassium also helps with muscle function and heart rhythm and, in later years, may reduce the risk of kidney stones and osteoporosis. An infant’s body keeps a steady amount of potassium in the bloodstream while excreting excess amounts through urine. The normal amount of potassium a baby of age of 0 - 6 months may require per day is 400.0 mg (Brannagan, 2019, Health Supplements Nutritional Guide, 2017). Calcium (Ca) is a nutrient that builds strong bones. The child gets a chance to build strong bones when in adolescence or teens. Children who get enough calcium start their adult lives with the strongest bones possible. This helps protect against bone loss later in life. Calcium also keeps the nerves and muscles functioning and plays a role in keeping the heart healthy (KidsHealth, 2017). Young children and babies need calcium and vitamin D to prevent a disease called rickets. Rickets softens the bones and causes bow legs, stunted growth, and sometimes sore or weak muscles. The normal amount of potassium a baby of age of 0 - 6 months may require per day is 200mg (KidsHealth, 2017; Food and Nutrition, 2001).

Breastfeeding is crucial in Africa, and considered the most cost-effective lifesaving intervention with the potential to reduce 13% of annual death among children under five (Turin & Ochoa, 2014). The purpose of this study was to examine seven breast-milk mineral and trace element (Sodium, Magnesium, Potassium, Calcium, Manganese, Copper and Iodine) concentrations from exclusively lactating mothers using the Neutron Activation Analysis, NAA and the Milk. Epithermal Instrumental NAA technique (EINAA).

## Materials and Methods

The study is a quantitative research with the aim determining the concentration of seven elements in the breastmilk of mothers from 23 communities in the Central Region of Ghana. Two main health facilities were used due to their accessibility and high utilization by patients. The selected facilities are located in Cape Coast and Elmina – and they provide both static and outreach child welfare services which captures mothers from towns in and around them. Cape Coast and Elmina are metropolis of the Central Region of Ghana, along the Gulf of Guinea (Figure 1). The original inhabitants of the towns and villages are mostly subsistence farmers and fishmongers (Vowotor *et al*, 2011). The volunteers’ occupations included petty traders, caterers, teachers, nurses, seamstresses, student, hairdressers, and some were unemployed.

**Figure 1:**
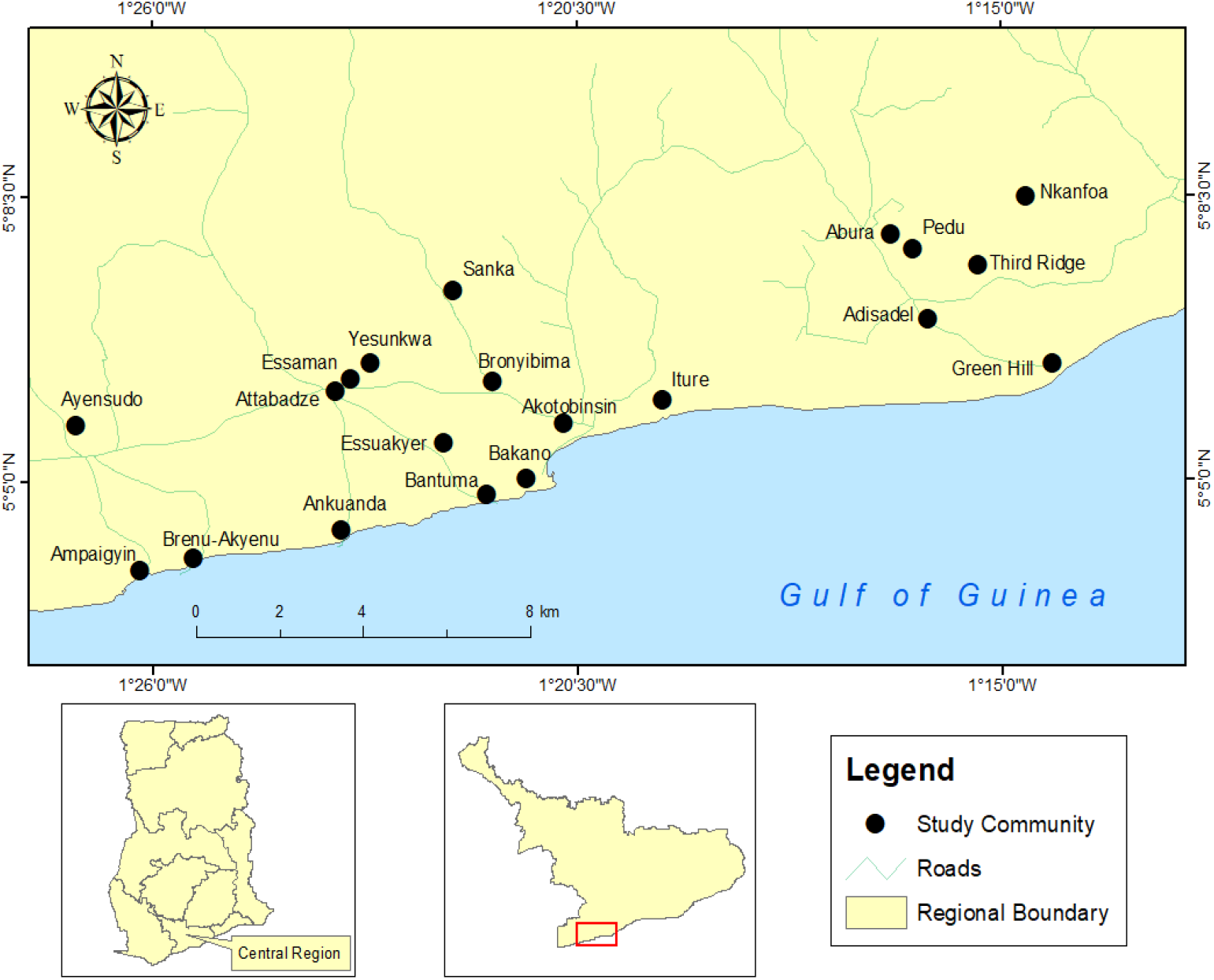
A map showing Cape Coast and its Environs, 2019.

### Study area

#### Sample collection

Data collection lasted for one month. The criteria for the selection was that the mothers should have volunteered to be part of the study, should have had full term pregnancies, practicing exclusive breastfeeding and was within the first six months of the babies’ lives. The study utilised the services of trained Midwife and Nurses who were service providers to these clients and were able to take breastmilk from those mothers aseptically. All of mothers were practicing exclusive breastfeeding within the first six months of the baby’s life.

Before the sample (breast milk) collection, the nipples and areolas of the breasts were cleaned with 70% methyl alcohol. The breast milk was collected by manual expression of approximately 10 – 20 mL and delivered directly into a sterile 60-mL plastic vials that is labelled with codes and immediately stored on ice, to prevent wastage, spoilage and contamination as recommended by BFN, 2009. The samples were transported to the storage point where they were put in a deep freezer with absolute temperature below – 20 °C. After the one month, the samples were transported in a vaccine carrier at the desired temperature to the Ghana Research Reactor-1 (GHARR-1) facility at the Ghana Atomic Energy Commission, Kwabenya, Accra for elemental analysis. Multivariate Analysis was used to analyse the results.

### Overview of Neutron Activation Analyses Technique Used

#### Epithermal Neutron Activation Analyses

Neutron Activation Analyses (NAA) is a sensitive analytical technique useful for performing both qualitative and quantitative multi-element analyses on samples. NAA offers sensitivities that are sometimes superior to those attainable by other methods, on the order of nano-gram level. NAA is accurate and reliable, generally recognized as the “referee method” of choice in the face other new procedures (Ali, 1999).

During NAA (Figure 3) a neutron interacts with the target nucleus (breastmilk) via a non-elastic collision, a compound nucleus forms which is now in an excited state. The excitation energy of the compound nucleus is due to the binding energy of the neutron with the nucleus. The compound nucleus will almost instantaneously de-excite into a more stable configuration through emission of one or more characteristic prompt gamma rays. In many cases, this new configuration yields a radioactive nucleus which also decays by emission of one or more characteristic delayed gamma rays, but at a much slower rate according to the unique half-life of the radioactive nucleus. Depending upon the particular radioactive species, half-lives can range from a fraction of a second to several years (Ali, 1999).

The basic essentials required to carry out an analysis of samples by NAA are a source of neutrons, instrumentation suitable for detecting gamma rays, and a detailed knowledge of the reactions that occur when neutrons interact with target nuclei. The sensitivities for NAA are dependent upon the irradiation parameters (i.e., neutron flux, irradiation and decay times), measurement conditions (i.e., measurement time, and detector efficiency); nuclear parameters of the elements being measured (i.e., isotope abundance, neutron cross-section, half-life, and gamma-ray abundance). Different types of reactors and different positions within a reactor can vary considerably with regard to neutron energy distributions and fluxes due to the materials used to moderate the primary fission neutrons. Most neutron energy distributions are quite broad and consist of three principal components: Thermal, Epi-thermal, and Fast. The thermal neutron component consists of low-energy neutrons (energies below 0.5 eV) in thermal equilibrium with atoms in the reactor’s moderator. The fast neutron component of the neutron spectrum (energies above 0.5 MeV) consists of the primary fission yielding neutrons which still have much of their original energy following Fission. The application of purely instrumental procedures is commonly called instrumental neutron activation analysis (INAA) (Ali, 1999). However, INAA methods using reactor flux neutrons for low level iodine measurement in biological materials suffer from high background activities from the activation products of major elements like sodium (Na), chlorine (Cl), manganese (Mn), potassium (K), bromine (Br) and Aluminum (Al) in the sample (Acharya & Chatt, 2009).

For many elements, the variation of the cross-section is inversely proportional to the neutron velocity (the l/ν law) and these are strongly activated by slow (i.e. thermal) neutrons. In contrast, other elements possess resonance cross-sections in the epithermal region, which are usually larger than the cross-sections by several orders of magnitude. Therefore, if a sample is irradiated with epithermal neutrons, it is expected that the activation yield of the “resonance” elements will be enhanced relative to those interfering nuclides which are activated mainly by thermal neutrons (Chisela, Gawlik & Bratter, 1986).

The NAA technique that employs only epi-thermal neutrons to induce (n, gamma) reactions by irradiating the samples being analyzed inside either cadmium or boron shields is called epi-thermal neutron activation analysis (ENAA). The epi-thermal neutron component consists of neutrons (energies from 0.5 eV to about 0.5 MeV) which is only partially moderated. In a typical unshielded reactor irradiation position, the epi-thermal neutron flux represents about 2% the total neutron flux (Ehmann & Vance, 1991). In reactor activation, this ENAA technique is performed by enclosing samples in thermal neutron filters such as cadmium or boron, which remove thermal neutrons from the reactor neutron spectrum. It has been applied to a variety of sample matrices including geological and biological materials (Chisela *et al*, 1986).

The EINAA methods are based on the fact that the resonance integral (I_0_) to thermal neutron (n,γ) cross-section (σ_0_) ratio (Q_0_) for ^127^I (24.8) is much larger than some of the interfering elements such as ^23^Na (0.59), ^37^Cl (0.69), ^27^Al (0.71), ^41^K (0.97), and ^55^Mn (1.053). The determination of iodine is sometimes hindered by EINAA using a cadmium (Cd) filter in cases of high dead-time from the neutron irradiated samples. Additionally, the background in the region of the 443-keV photopeak of ^128^I is often dominated by Compton background from the γ-rays of ^24^Na, ^56^Mn, ^38^Cl, ^42^K, ^28^Al and ^82^Br. The combination of EINAA using cadmium and/or boron shields and anticoincidence gamma-ray spectrometry (AC) is used to suppress the background, which in turn helps to improve the detection limit of these elements (Acharya, 2009).

### Sample Preparation and Irradiation

500 mg of each sample was weighed into a smaller polyethylene vials via the micrometre pipette (Figure 2), after shaking them in their original containers to ensure uniformity. The polyethylene capsule of diameter 1.2 cm and height 2.35 cm containing the liquid samples were in turn put in to bigger polyethylene capsule of diameter 1.6 cm and height 5.5 cm (Rabbit capsule), that is Double encapsulation and smoothly heat sealed with a soldering rod. The irradiation vials (capsules) that were used were pre-cleaned by washing them first with distilled water and then soaked in an acidic reagent for 24 hrs, and then rinsed in distilled deionised water. The irradiation vials were further soaked in nitric acid (HNO_3)_ for another 24 hrs. They were then rinsed thoroughly with distilled deionised water and air-dried in a clean fume hood.

**Figure 2:**
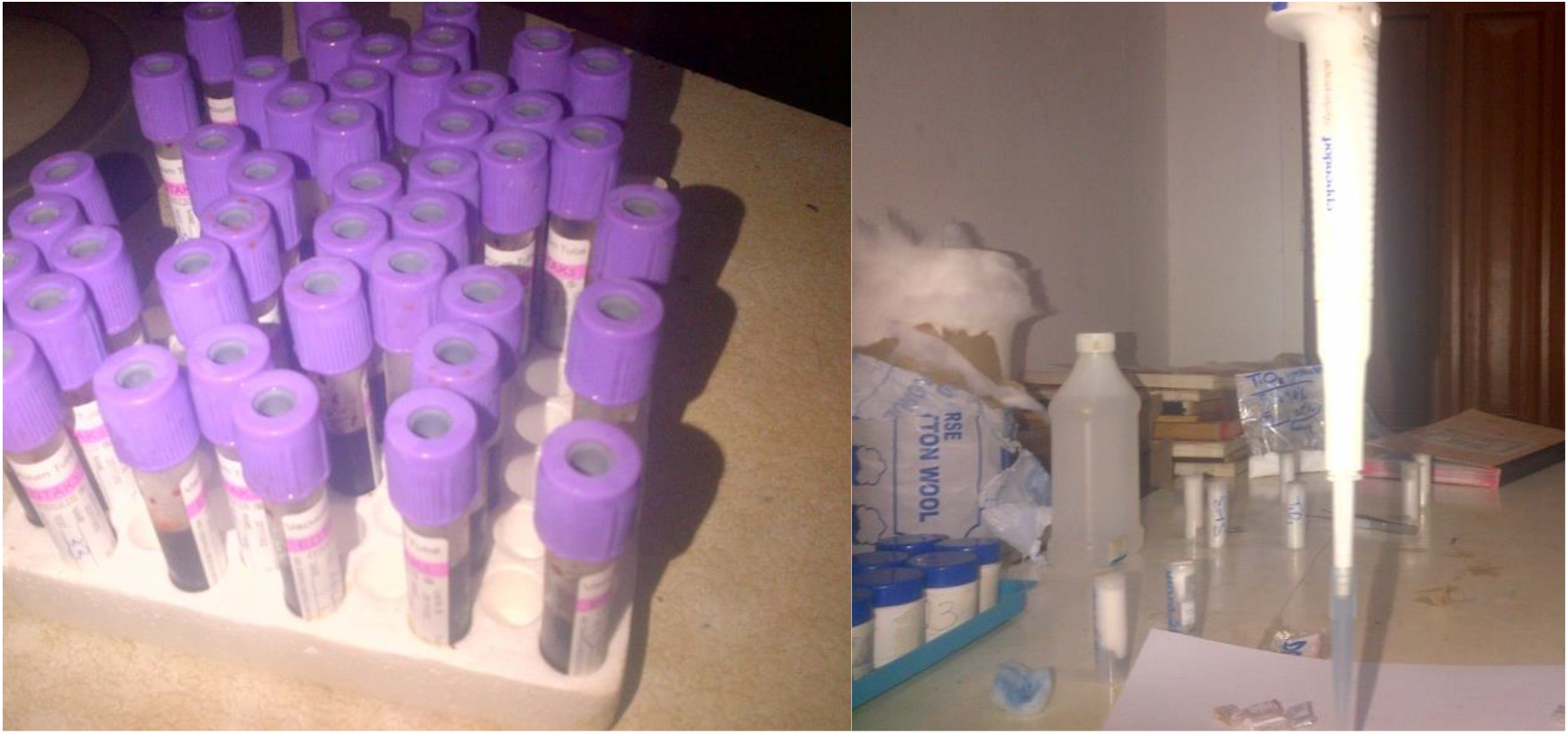
Breast Milk Samples Being Prepared for Irradiation at GHARR-1.

**Figure 3:**
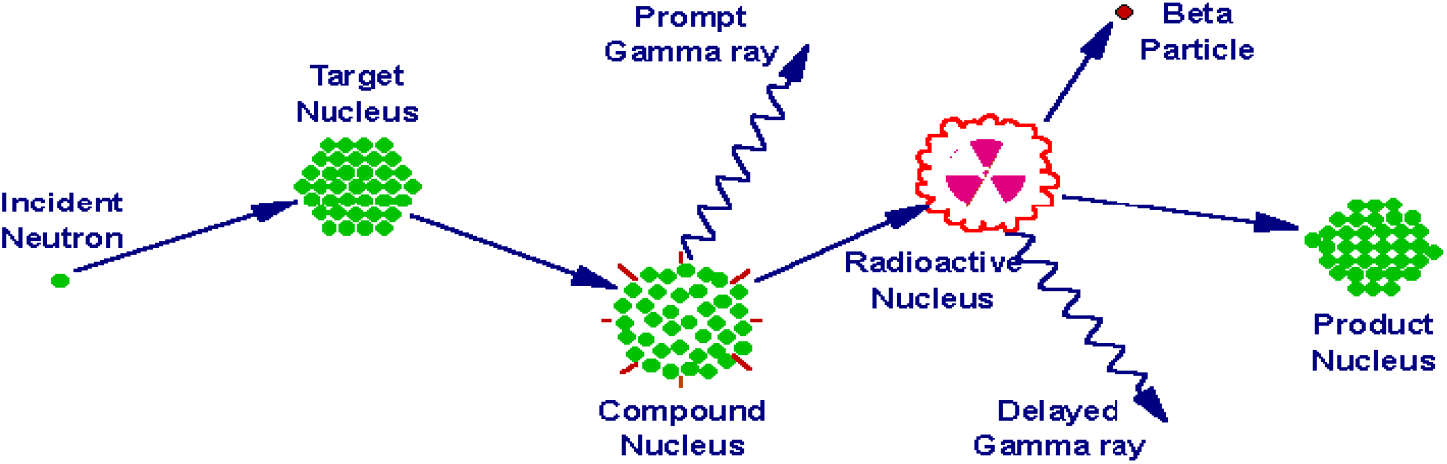
A process of neutron capture followed by emission of gamma ray (Glascock, 2003)

**Figure 4:**
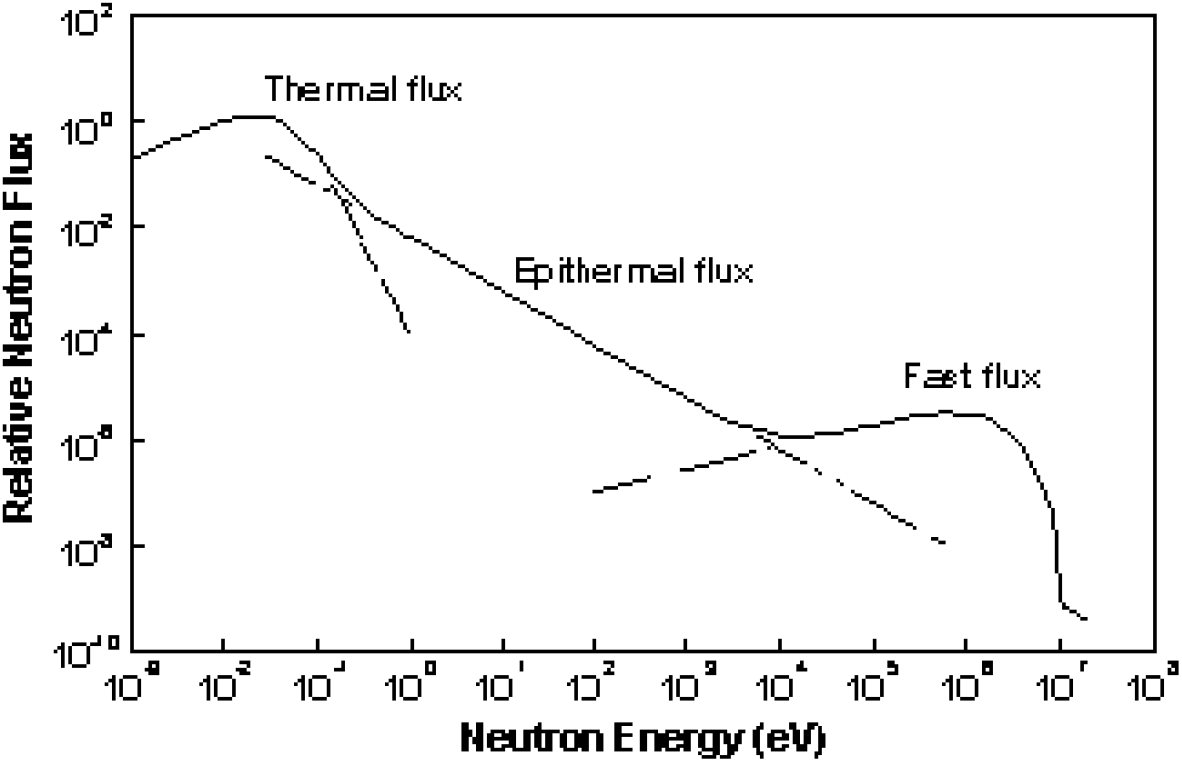
A typical reactor neutron energy spectrum showing the various components used to describe the neutron energy regions (Glascock, 2003).

**Figure 5:**
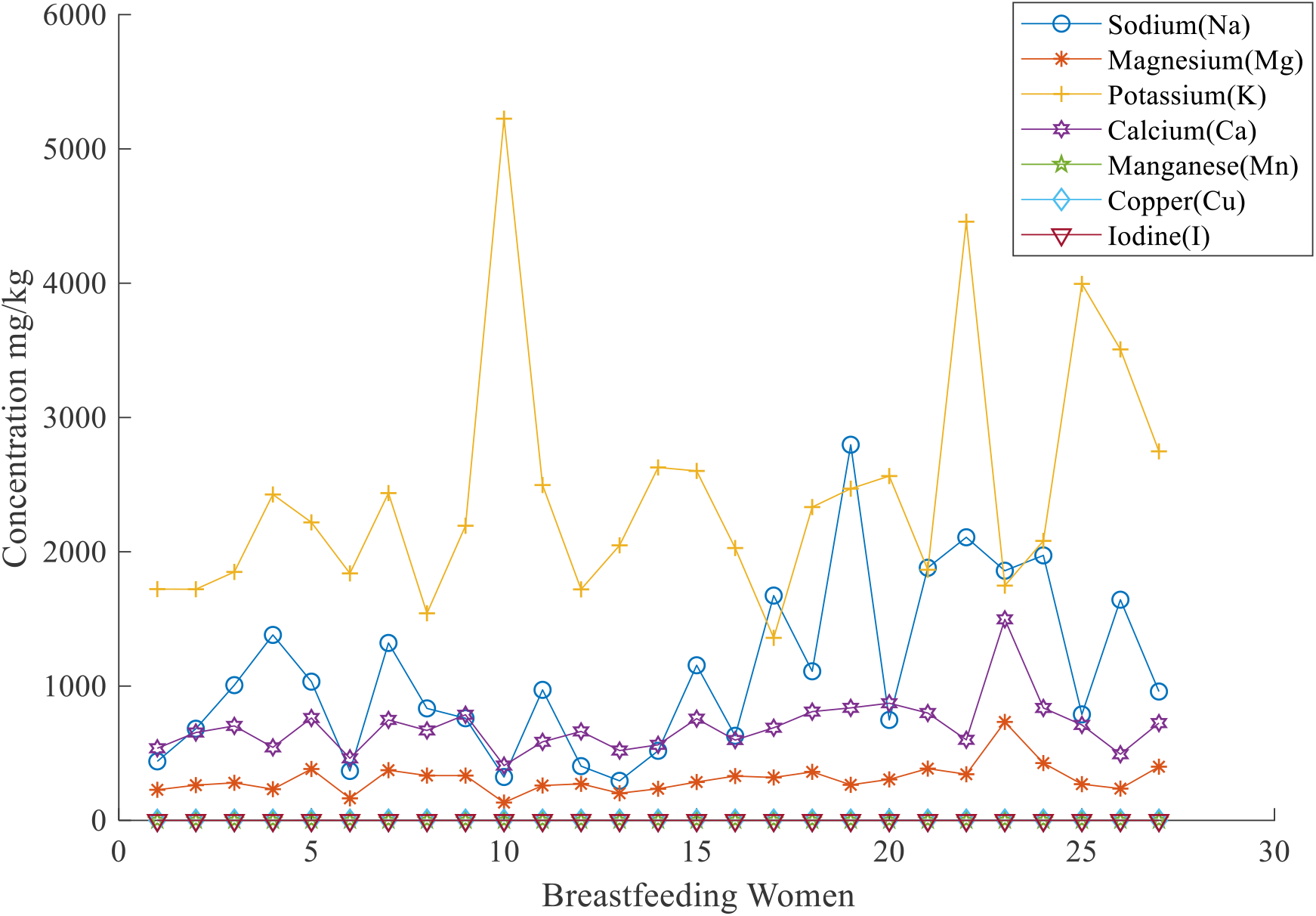
Concentration of the seven elements in the breastfeeding mothers.

To validate the procedure, various of standard reference materials namely IAEA-336 (trace and minor elements in Lichen), IAEA-407 (trace elements and methyl mercury in fish tissue), IAEA-350 (trace elements in tuna fish homogenate) and SRM 1577b (Bovine liver) were prepared and packed similarly as the samples (IAEA, 1999; IAEA, 2003; EVISA1, 2010; EVISA2, 2010). Samples were transferred into irradiation sites through pneumatic transfer system at a pressure of 60 psi. The irradiation was categorized according to the half-life of the element of interest. Samples and controls were irradiated in the Ghana Research Reactor (GHARR-1) at the Ghana Atomic Energy Commission (GAEC), operating at 15 KW at a thermal flux of 5×10^11^ ncm^-2^s^-1^. Three millimeters (3mm) thick of flexible boron was used to cut off thermal neutrons in order to assess epithermal neutrons. 94.7% accuracy was achieved.

### Relative Standardization

In the relative standardization method, a chemical standard (index std) of known mass, Wstd, of the element is co-irradiated with the sample of unknown mass Wsam. When the samples to be irradiated is short-lived radionuclide, both the standard and sample are irradiated separately under the same conditions, usually with a monitor of the same neutron fluence rate and both are counted in the same geometrical arrangements with respect to the gamma-ray energy. It is then assumed that the neutron flux, cross section, irradiation times and all other variables associated with counting are constant for the standard and the sample at a particular sample-to-detector geometry. With this assumption, the neutron activation equation then reduces to:

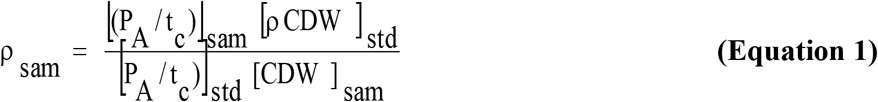

where (P_A_/t_c_)std and (P_A_/t_c_)sam are the counting rates for the standard and sample respectively, ρ _*std*_ and ρ _*sam*_ are the counting concentrations of the standard and the element of interest respectively, C_std_ and Cs_am_ are the counting factors for standard and sample, D_std_ and D_sam_ are the decay factors for the standard and sample respectively.

### Qualitative and Quantitative Analysis

The qualitative analysis involves the determination of the seven elements in the breast milk samples by the identification of the spectra peaks and assigning corresponding radionuclides and hence the elements present. The quantitative analysis, involves the calculation of the areas under the peaks of the identified elements and converting them into concentrations using an appropriate software or equation(s) (Alfassi, 1994). The counting of the induced radioactivity was performed by a PC-based γ-ray spectrometry. It consists of an n-type high purity Germanium (HPGe) detector (model GR2518) coupled to a computer-based Multichannel Analyzer via electronic modules and a spectroscopy amplifier (model 2020, Canberra Industries Incorporated). The relative efficiency of the detector is 25% with an energy resolution of 1.8 KeV at γ-ray energy of 1332 KeV of ^60^Co. The qualitative analysis was achieved by means of ORTEC EMCAPLUS Multichannel Analyzer (MCA) Emulation software. A Microsoft Window–based software, MAESTRO, was used for spectrum analysis (Serfor-Armah, Carboo, Akuamoah, & Chellube, 2018). This software identifies the various photo peaks, estimates and works out the areas under them. The other quantitative measurements were done using the concentration equation (Equation 1) in a Microsoft Excel programme for calculating the elemental concentrations in μg/g. The detection limit (DL) of the various elements of interest for Neutron Activation Analysis and the nuclear data have been summarized are shown Table 1.

**Table 1:**
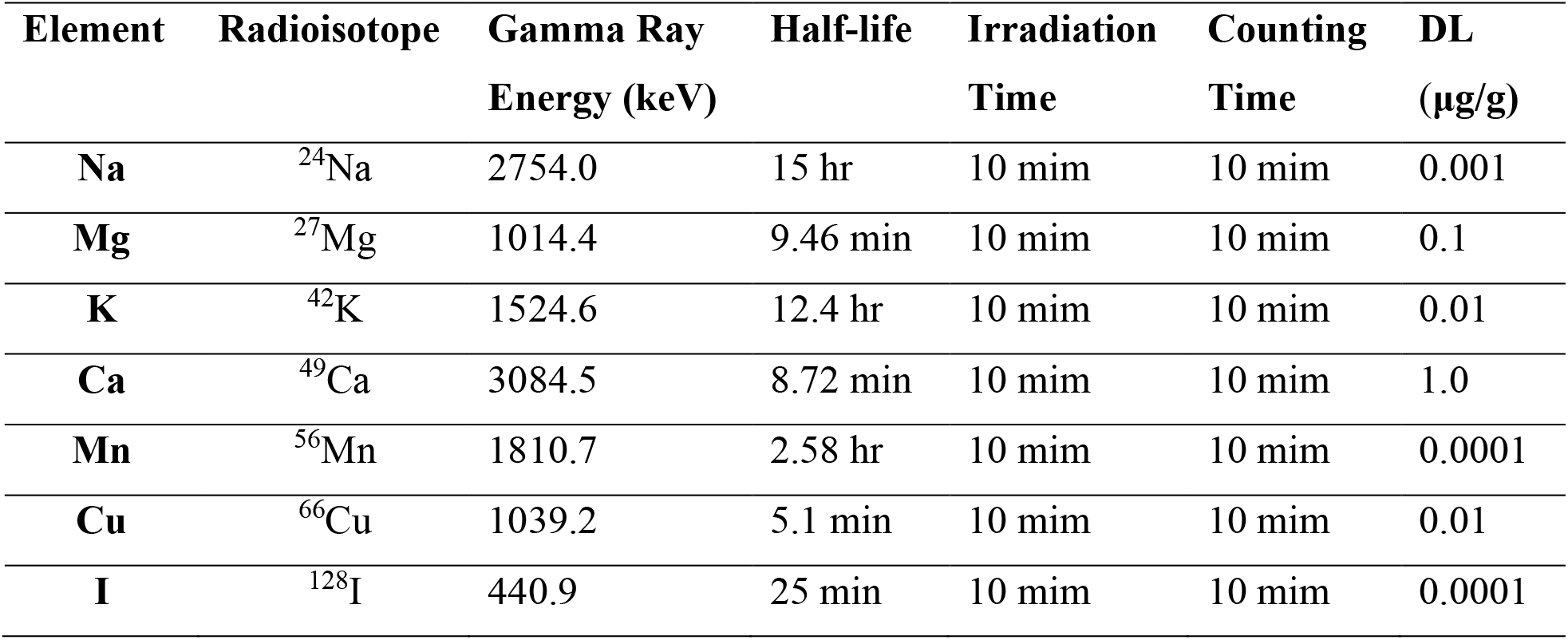
Nuclear Data of the Seven Elements.

## Results

### Assessment According to Concentrations

The concentration of each micro nutrient investigated in the breast milk is as shown in Table 2. Nutrient concentrations ranged within the following intervals: Na: 294.4-2797.0 mg/kg; Mg: 132.4-426.7 mg/kg; K: 1359.0-5225.0 mg/kg; Ca: 408.1-1497.0 mg/kg; Mn: 0.0018-1.115 mg/kg; Cu: 0.2235-1.0199 mg/kg; and I: 0.029-0.1445 mg/kg, and their mean concentrations were: Na: 1098.6259 mg/kg; Mg: 309.2481 mg/kg; K: 2438.0741 mg/kg; Ca: 697.4037 mg/kg; Mn: 0.4028 mg/kg; Cu: 0.6944 mg/kg; and I: 0.0613 mg/kg. Arranging the micro nutrients in terms of abundance in the breast milk yielded the order K > Na > Ca > Mg > Cu > Mn > I.

**Table 2:** Summary of Breast Milk Concentrations of Lactating Mothers.

|  | Sodium<br>(Na) | Magnesium<br>(Mg) | Potassium<br>(K) | Calcium<br>(Ca) | Manganese<br>(Mn) | Copper<br>(Cu) | Iodine<br>(I) |
| --- | --- | --- | --- | --- | --- | --- | --- |
| Min | 294.4000 | 132.4000 | 1359.0000 | 408.1000 | 0.0018 | 0.2235 | 0.0290 |
| Max | 2797.0000 | 426.7000 | 5225.0000 | 1497.0000 | 1.1150 | 1.0199 | 0.1445 |
| Mean | 1098.6259 | 309.2481 | 2438.0741 | 697.4037 | 0.4028 | 0.6944 | 0.0613 |
| SD | 36.4403 | 23.7186 | 176.349 | 35.2538 | 0.0384 | 0.0079 | 0.0102 |

### Assessment According to Recommended Dietary Allowable

Recommended Dietary Allowable (RDA) is the dietary requirement for a micronutrient. It is an intake level which meets a specific criterion for adequacy, thereby minimizing risk of nutrient deficit or excess. (Dietary Reference Intakes, 2013). Exclusively breastfed babies take in an average of 25 oz (750 ml) or 708.738 g per day between the ages of 1 month and 6 months (Bonyata, 2019). If the concentration of element in milk per day (RDA) is expressed in mg/d and the average intake of milk per day be represented by M g/d, then the concentration of elements in the breast milk represented by N mg/g of sample can then be given by the relation expressed in equation 2 (Vowotor *et al*, 2011);

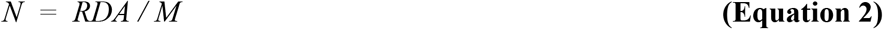

The concentration of the elements investigated is therefore calculated as shown in Table 3. Also, on Table 3 are the Average Daily Intake, ADI, calculated using equation 3, from the Mean Measured Concentrations, MMC.

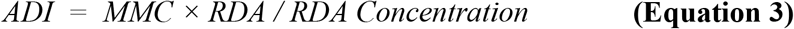

**Table 3:** RDA (mg/kg) and their Corresponding Concentrations (mg/kg) of Elements for Babies below 6 Months.

| Element | Na | Mg | K | Ca | Mn | Cu | I |
| --- | --- | --- | --- | --- | --- | --- | --- |
| **RDA** | 120.0 | 30.0 | 400.0 | 200.0 | 0.003 | 0.2 | 0.09 |
| **RDA Concentration** | 169.3 | 42.3 | 563.4 | 281.7 | 0.0042 | 0.28 | 0.13 |
| **Mean Measured Concentration** | 1098.6259 | 309.2481 | 2438.0741 | 697.4037 | 0.4028 | 0.6944 | 0.0613 |
| **Average Daily Intake from Concentration** | 778.71 | 219.32 | 1730.97 | 495.14 | 0.29 | 0.50 | 0.04 |
| **Difference Between the RDA & Measured RDA** | + 658.71 | + 189.32 | + 1330.97 | + 295.14 | + 0.287 | + 0.3 | – 0.05 |
| **HI** | **6.49** | **7.31** | **4.33** | **2.48** | **95.90** | **2.48** | 0.47 |

RDA Values – (Food and Nutrition, 2001, Mrunal, 2018)

### Except I

The elemental concentrations of all the elements measured were found to be very high compared to concentrations calculated from the RDA (Table 3). In a decreasing order of their mean concentrations, the elements investigated can be arranged as follows: K > Na > Ca > Mg > Cu > Mn > I. Looking at difference between the RDA values and the measured/calculated RDA values, the negative sign (–) on the I value denotes a value below the recommended RDA, whiles the positive sign (+) denotes values are above the recommended RDA.

### Assessment According to Health Risk Estimation (HI)

To estimate the health effects, hazard index (HI), the estimated lifetime average daily dose of each chemical is compared to its Reference Dose (RfD). The reference dose represents an estimate of a daily consumption level that is likely to be without deleterious effects in a lifetime. Based on the equation detailed in the US. EPA handbook (Laar, Fianko, Akiti, Osae & Brimah, 2011):

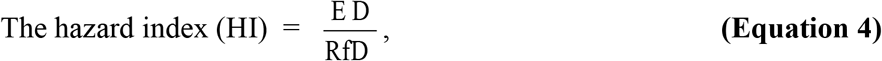

Where, ED = Estimated Dose and RfD = Reference Dose

Pearson correlation was calculated and presented on Table 5 showed normal distribution in concentrations of the elements in the fish consumed. The variables are continuous and linear in relationship between the variables. Though 95% confidence level was used to ascertain the strength of their relationship, there are other strongly correlated elements with high coefficients of determination, hence cannot be ruled out and the focus should be on strength of relationship and the amount of variance shared while reporting statistical significance (Pallant, 2007).

Arranging the micro nutrients in terms of abundance in the breast milk yielded K > Na > Ca > Mg > Cu > Mn > I. The low standard deviation (SD) values obtained indicated that the spatial distribution of the individual micro nutrients in their breast milk was uniform. So, for this age group in except I, all the levels of K, Na, Ca, Mg, Cu and Mn is far above the recommended daily dose. Table 4 gives suggestions of likely adverse health effects for these elements for the group. The estimated health risk associated with these breastmilk consumptions is also presented on Table 3. HI < 1 suggests an unlikely adverse health effects whereas HI > 1 suggests the probability of adverse health effects (Laar, Fianko, Akiti, Osae and Brimah, 2011). High HI values for the trace elements investigated that registered values > 1 have been highlighted. The HI values calculated on Table 3 have buttressed the trend in the assessment according to the concentrations and the recommended dietary allowable. So, their trends as depicted on Table 4 would help give suggestions of likely adverse health effects for these children in later life.

**Table 4:**
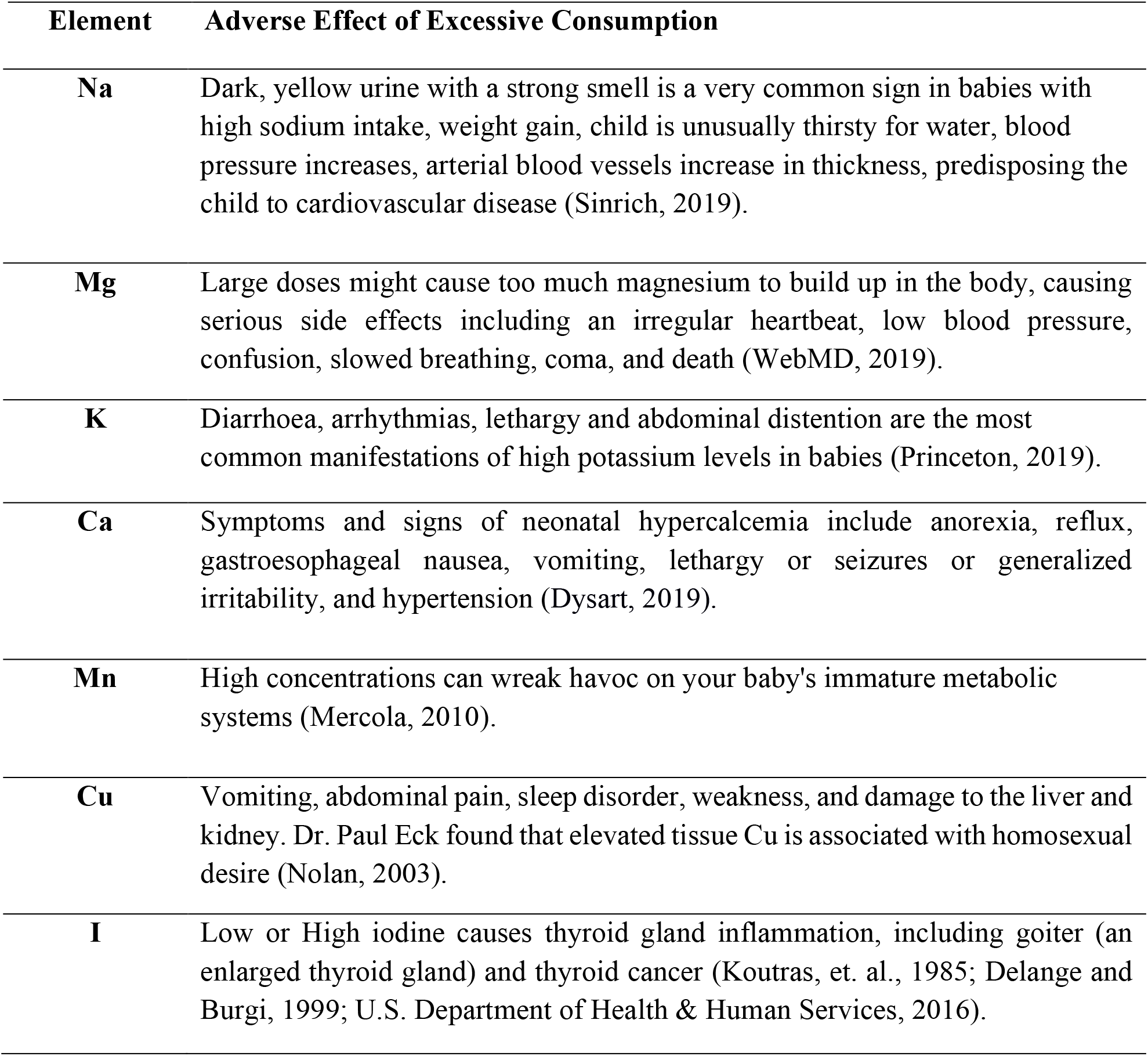
The Seven Elements and the effect of its excess.

| Element | Adverse Effect of Excessive Consumption |
| --- | --- |
| **Na** | Dark, yellow urine with a strong smell is a very common sign in babies with high sodium intake, weight gain, child is unusually thirsty for water, blood pressure increases, arterial blood vessels increase in thickness, predisposing the child to cardiovascular disease (Sinrich, 2019). |
| **Mg** | Large doses might cause too much magnesium to build up in the body, causing serious side effects including an irregular heartbeat, low blood pressure, confusion, slowed breathing, coma, and death (WebMD, 2019). |
| **K** | Diarrhoea, arrhythmias, lethargy and abdominal distention are the most common manifestations of high potassium levels in babies (Princeton, 2019). |
| **Ca** | Symptoms and signs of neonatal hypercalcemia include anorexia, reflux, gastroesophageal nausea, vomiting, lethargy or seizures or generalized irritability, and hypertension (Dysart, 2019). |
| **Mn** | High concentrations can wreak havoc on your baby's immature metabolic systems (Mercola, 2010). |
| **Cu** | Vomiting, abdominal pain, sleep disorder, weakness, and damage to the liver and kidney. Dr. Paul Eck found that elevated tissue Cu is associated with homosexual desire (Nolan, 2003). |
| **I** | Low or High iodine causes thyroid gland inflammation, including goiter (an enlarged thyroid gland) and thyroid cancer (Koutras, et. al., 1985; Delange and Burgi, 1999; U.S. Department of Health & Human Services, 2016). |

## Discussion

### Assessment According to Correlation Coefficient and Sources of Trace Elements

An important source of nutrients or trace elements is from the food, processed water and groundwater discharges (Kelly and Moran, 2002). Accordingly, it is important to characterize each distinct source of trace elements and determine its contribution to baby and mother. Correlation coefficients between the seven elements and their respective significance at 95% significance level in the breast milk are in Table 5. The interpretation of the strength of the correlation coefficients usually depends on the researcher as suggested by the guidelines from Rumsey 2010. In this study, values between ± 0.500≤ r ≤ ±0.999 are considered strong correlations. The level of statistical significance that does not indicate how strongly two variables are associated (is given by rho), but instead it indicates how much confidence we should have in the results obtained. Because the significance of rho is strongly influenced by the sample size, smaller sample sizes does not reach statistical significance as compared to larger sample sizes which may even be statistically significant at small (weak) correlations (Pallant, 2007). It was found out that even though 4 correlations were strong, 3 were statistically significant. This means that we can exude 95% confidence that the correlation between Ca and Mg is strongly correlated with a coefficient of 0.905. As expected, there was a moderately strong correlation between Na and Mg since main source of Na is added salt. It is worth noting that salt is one of cheapest essential exporting commodities from the study area. Salt is mined at Elmina and sold in all the communities in the Central Region. Most of the foods eaten by participants are seasoned with salt. For example, some mothers said they eat bread which are made from wheat rich in Mg and salt for breakfast. Other salted foods rich in Mg are the soups made from vegetables, seafood (salmon, mackerel, tuna) and beans. Salted roasted peanuts with banana are a delicacy in Ghana.

**Table 5:** Correlation coefficients of the concentration of the seven elements in breast milk.

|  | **Na** | **Mg** | **K** | **Ca** | **Mn** | **Cu** | **I** |
| --- | --- | --- | --- | --- | --- | --- | --- |
| **Na** | 1 |  |  |  |  |  |  |
| **Mg** | **0.465\*** | 1 |  |  |  |  |  |
| **r<sup>2</sup>** | 0.014 |  |  |  |  |  |  |
| **K** | 0.035 | -0.284 | 1 |  |  |  |  |
| **r<sup>2</sup>** | **0.861** | 0.152 |  |  |  |  |  |
| **Ca** | **0.459\*** | **0.905\*\*** | -0.304 | 1 |  |  |  |
| **r<sup>2</sup>** | 0.016 | 0.000 | 0.124 |  |  |  |  |
| **Mn** | 0.250 | **0.836\*\*** | -0.214 | **0.733\*\*** | 1 |  |  |
| **r<sup>2</sup>** | 0.208 | 0.000 | 0.285 | 0.000 |  |  |  |
| **Cu** | **0.832\*\*** | **0.545\*\*** | 0.192 | **0.531\*\*** | **0.389\*** | 1 |  |
| **r<sup>2</sup>** | 0.000 | 0.003 | 0.337 | 0.004 | 0.045 |  |  |
| **I** | -0.171 | 0.002 | -0.175 | 0.020 | -0.371 | -0.269 | 1 |
| **r<sup>2</sup>** | 0.393 | **0.993** | 0.384 | **0.920** | 0.057 | 0.174 |  |

\*Correlation is significant at the 0.05 level (2-tailed).

There is a strong correlation of 0.905 between Na and Ca. This can also be attributed to salted foods rich in Ca such as sardine, salmon, beans and leafy greens which are found in most mothers’ daily diets like Yam and palava sauce, Fufu and soup, and Gari and beans. Chocolate which is rich in Cu is made from Cocoa. Ghanaians drink a lot of cocoa product such as chocolates which are spiced with salt therefore can be the source of correlation between Na and Cu. Other Cu rich foods like lobsters, squid, oysters and organ meats such as liver and kidneys are also seasoned with salt before used for soup. Mg has strong correlation with Ca, Mn and Cu of 0.905, 0.836 and 0.545 respectively. Mg and Ca rich foods like the legumes and seafoods may be another source such as *Gari* and Beans or soups are made from seafood.

Mn combined with other elements is widely distributed in Earth’s crust. Manganese is second only to iron among the transition elements in its abundance in Earth’s crust; it is roughly similar to iron in its physical and chemical properties but harder and more brittle. It occurs in a number of substantial deposits, of which the most important ores (which are mainly oxides) consist primarily of manganese dioxide (MnO_2_) in the form of pyrolusite, romanechite, and wad. Manganese is essential to plant growth and is involved in the reduction of nitrates in green plants and algae. It is an essential trace element in higher animals and participates in the action of many enzymes. Lack of manganese causes testicular atrophy while excess is toxic (Encyclopaedia Britannica. 2014). It is not surprising to find Mn also exhibiting a strong correlation with Ca and Cu of about 0.733 and 0.389 respectively. Foods like beans and spinach are rich and a good source of Mn and Ca. seafood such as Oysters, Lobster, Squid are also rich in Cu are usually made into stews a delicacy eaten with brown rice also rich in Mn are a delicacy for participants. The last good correlation is between Ca and Cu of 0.531 may probably come from green leafy vegetables such as broccoli, parsley, spinach, and lettuce, Cabbage, *Kontonmire* etc as their main source since they are used in making nutritious sauces.

## Conclusion

Diseases affecting people in later years of their lives are precipitant from childhood through what was eaten and the pattern outlines by their parents either out of curiosity, ignorance or lack of education on nutrition. From Table 3 only one element was found to be in the normal RDA range the rest very high. Mn was found to be the element exceeding the DRA threshold and in very high quantity consumed by these babies without their mother’s knowledge. These children could be exposed to metabolic disorders or unexplained diseases in future without knowing where they originated from. Because Na is in abundance, it is incorporated intentionally or unintentionally into every food by the locals. Na as found in salt is used in all things that are edible so accumulation of it starts right from childhood. This could also lead to the unexplained hypertension in early adulthood.

## Data Availability

Datasets used and/or analysed during the study are available from the corresponding author (IKA) on reasonable request.

## Acknowledgment

The authors wish to express their sincere appreciation to the Ghana Health Service (GHS), the directors of the 2 Health facilities for allowing the study to be carried out. Appreciation also go to the mothers, nurses and technicians who collected data and the staff of Ghana Atomic Energy Commission at Kwabenya-Accra, for their assistance in running the tests and analysing of the samples.

## Authors contributions

Michael Dela Vowotor is the main author of the manuscript together with Irene Korkoi Aboh and Andrews Adjei Druye. He was responsible for the conception, study design, data collection, data analysis, Irene Korkoi Aboh is responsible for the drafting of the manuscript. Andrews Adjei Druye was responsible for content validity of the manuscript.

## Ethical approval and consent to participate

The study was approved by the Ghana Health Service Ethical Review Committee with number GHS-ERC-02/05/15. Voluntary participation of participants was accorded with respect, dignity and confidentiality because samples taken were a part of their body issue.

## Conflict of interest

The authors declare that they have no competing interests.

## Notes

### Competing Interest Statement

The authors have declared no competing interest.

### Clinical Trial

This study is not a clinical trial - so it was not registered

### Funding Statement

There was no funding for this work

### Author Declarations

Letter of ethical approval from the Ghana Health Service Ethical Review Committee with number GHS-ERC-02/05/15.

